# A multivariate geostatistical framework for combining multiple indices of abundance for disease vectors and reservoirs: A case study of *rattiness* in a low-income urban Brazilian community

**DOI:** 10.1101/2020.07.31.20165753

**Authors:** Max T. Eyre, Ticiana S. A. Carvalho-Pereira, Fábio N. Souza, Hussein Khalil, Kathryn P. Hacker, Soledad Serrano, Joshua P. Taylor, Mitermayer G. Reis, Albert I. Ko, Mike Begon, Peter J. Diggle, Federico Costa, Emanuele Giorgi

**Author notes:** These authors contributed equally to this work (Federico Costa, Emanuele Giorgi).

## Abstract

A key requirement in studies of endemic vector-borne or zoonotic disease is an estimate of the spatial variation in vector or reservoir host abundance. For many vector species, multiple indices of abundance are available, but current approaches to choosing between or combining these indices do not fully exploit the potential inferential benefits that might accrue from modelling their joint spatial distribution. Here, we develop a class of multivariate generalized linear geostatistical models for multiple indices of abundance. We illustrate this novel methodology with a case study on Norway rats in a low-income urban Brazilian community, where rat abundance is a likely risk-factor for human leptospirosis. We combine three indices of rat abundance to draw predictive inferences on a spatially continuous latent process, *rattiness*, that acts as a proxy for abundance. We show how to explore the association between *rattiness* and spatially varying environmental factors, evaluate the relative importance of each of the three contributing indices, assess the presence of residual, unexplained spatial variation, and identify *rattiness* hotspots. The proposed methodology is applicable more generally as a tool for understanding the role of vector or reservoir host abundance in predicting spatial variation in the risk of human disease.

## 1 Introduction

In studies of endemic vector-borne and zoonotic diseases, estimates of vector and reservoir host abundance, including spatial variation in abundance, are often needed to inform predictive models of disease risk and to guide the decision-making process for the implementation, monitoring and evaluation of control programmes [1]. Detecting all members of a target population at a sampled location is impossible for most disease vector or reservoir species. Consequently, indirect methods of determination are often used in ecological studies to obtain indices that quantify relative abundance [2–4]. Here, since our focus is on the effect of vector and reservoir host populations on human health, we use the term “abundance” loosely to denote all ecological processes that are associated with animal abundance, for example animal presence and activity, and that can be used to quantify exposure, including spatial variation in exposure, to a disease of interest.

In the absence of a gold-standard index of animal abundance, many different indices are commonly used for a single species, sometimes within the same study. For example, in the case of rodents, indices derived from traps, camera traps, counts (of animals, tracks, burrows and faeces), track plates and gnawing pegs have all been used to estimate rat abundance [2, 5, 6]. Similarly, for insects, a wide range of entomological indices are used. For example, occurrence, density (per unit time, surface area, person or other sampling unit), human-biting rates and the human blood index are used to estimate adult mosquito abundance [7–10]. When data for multiple imperfect indices of abundance has been collected within a study area, methods which can jointly model these quantities may improve prediction and inference. Such methods are also useful when different traps (or protocols) with different detection probabilities and biases are used to collect data for the same index [11, 12]. However, current approaches to combining multiple indices of abundance do not exploit the inferential benefits that might accrue from their joint modelling.

Many recent studies have attempted to model the spatial distribution of disease vectors, for example in the context of malaria [12], dengue [13], Chagas disease [14], Human African trypanosomiasis [15], schistosomiasis [16], leishmaniasis [17], West Nile virus [18] and rodent-borne zoonoses such as leptospirosis [19], plague [20], hantavirus [21] and *Bartonella* spp. [22]. In such cases, direct determination of vector or reservoir host abundance throughout the study area is often impractical, because of the extensive sampling effort required. A practical solution is to sample a finite set of locations and use statistical modelling to make predictions at unobserved locations by capturing spatial correlation and associations with environmental drivers of abundance. Here, we achieve this by the use of model-based geostatistics [23, 24], a branch of spatial statistics that provides a principled likelihood-based approach for mapping of geo-referenced outcomes. A geostatistical model is an extension of a generalised linear mixed model that accounts for covariate effects and otherwise unexplained spatial variation in the outcome of interest. Geostatistics has been used in a range of scientific disciplines, including ecology [25, 26] and epidemiology [24].

In this study, therefore, having described our motivating application in Section 1.1, in Section 2.1 we set out statistical criteria for combining multiple indices of vector and reservoir host abundance and review the literature for existing and relevant methodologies. Then, in Sections 2.2 to 2.4 we present a new class of multivariate generalized linear geostatistical models for combining multiple indices of abundance, which exploit the spatial correlation both within and across indices. In Section 3, we illustrate the development and application of the methodology in the context of a case study on the Norway rat, a reservoir for infectious diseases in low-income urban communities in Salvador, Brazil. Mapping Norway rat abundance is essential for investigating its role in disease transmission and developing more targeted rodent control strategies. In Section 4, we apply our novel methodology to the analysis of data collected for three indices of rat abundance, make inferences about the association of environmental variables with *rattiness*, our proxy for rat abundance, and map *rattiness* for the entire study area. We then assess the relative contribution of each index to the spatial predictions. Finally, in Section 5 we discuss the strengths, limitations and wider applicability of the developed methodology.

### 1.1 Motivating application: Mapping the abundance of Norway rats, a reservoir for *Leptospira* in urban Brazil

Leptospirosis is a widespread and neglected zoonotic disease caused by pathogenic bacteria from the *Leptospira* genus. It is among the leading zoonotic causes of morbidity and mortality globally, with more than one million human cases and 58,000 deaths reported each year [27, 28]. Humans are infected via direct contact with animal reservoirs or through contact with soil or water contaminated by bacteria shed in the urine of infected animals [29, 30]. In tropical low- and middle-income countries, including Brazil, low-income urban communities (often referred to as ‘informal settlements’ or ‘slum communities’ in the literature) are at increased risk for leptospirosis due to poor sanitary conditions, flooding, intense environmental contact and abundant local rat reservoir populations [31, 32].

Globally, the Norway rat, *Rattus norvegicus*, is a major reservoir host for *Leptospira* spp. and several other pathogens, and thrives in low-income peri-urban and urban environments where food and harbourage are plentiful [30, 33–37]. Norway rat populations have been found to have high prevalences of leptospiral infection in Brazil, Argentina, Japan and Canada [19, 30, 36, 38], and high daily leptospire shedding rates have been recorded in Salvador, Brazil [30].

Associations between the risk of leptospiral infection in humans and peri-domiciliary rat infestation [31, 32, 34, 35, 39] and rodent sightings [31, 32, 35] have been reported in multiple studies in Brazil. However, the link between rat population abundance and risk of spill-over infection to humans is poorly understood [32], partly due to limited knowledge about the distribution and abundance of rats within urban environments [6]. As a result, several ongoing eco-epidemiological studies in Salvador, Brazil, aim to address this knowledge gap and generate evidence about the impact of rat control measures on disease transmission [40] through the collection and analysis of human seroprevalence and rat abundance data.

However, estimation of rat abundance in complex urban settings is hindered by a lack of reliable measurement tools [2, 6]. In studies of the Norway rat in Salvador, a combination of rat trapping, surveys for signs of rodent infestation and track plates are routinely used as indices of relative abundance [6, 41]. Track plates are plastic plates which are coated in ink and placed on the ground to detect rat paw and tail markings. A recent study has shown that track plate measurements are correlated with those of rodent infestation surveys and rat trapping [6]. The use of alternative tools, such as track plates, which are cheaper and can be deployed faster and more easily, allows for a more cost-effective design of studies, while reducing the impact of the loss of sampling days and equipment due to violence (associated with drug-trafficking groups operating within these communities) and theft. However, different indices may have distinct biases and collecting data for multiple indices within a study site can allow for a richer and more comprehensive measurement of relative abundance. In this challenging context, statistical modelling can be especially useful in order to use of all the information collected from multiple measurement tools and deliver optimized inferences about rat abundance.

Our research question is: how should we develop a joint geostatistical model for multiple indices to map rat abundance? Our ultimate goal is to develop a reliable modelling approach that can be used to identify rat abundance hotspots to guide future investigations of environmental contamination and rodent control.

## 2 Model and methods

### 2.1 Developing an approach for combining multiple indices of abundance

To develop an objective and statistically principled approach for combining multiple indices of abundance, we propose that a statistical model should meet the following six criteria.

C1 It should account for the appropriate sampling distribution of each index through the use of a suitable conditional likelihood function.

C2 It should not require all indices to be taken at a common set of locations. C3 It should account for spatial correlation both within and across indices.

C4 It should allow for the prediction of abundance at all locations within the study area and quantify the uncertainty associated with those predictions.

C5 It should allow for the quantification of the relative contribution of each index to the spatial predictions. C6 It should allow for the incorporation of spatially referenced covariates.

We now review existing methodologies in the literature and assess how well they meet these criteria.

One of the simplest and most commonly used approaches is to directly combine data for multiple indices into a single index by averaging their values and modelling this using a standard linear regression model [42]; this approach violates criteria C1 to C3. A second approach is to model each index separately and independently using linear regression models, and to combine the resulting predictions [42]. Although this approach respects C1, it does not take advantage of the inferential benefits that would accrue from C3.

In a third approach, a composite index is created using a weighted combination of multiple indices. The weighting is often based on a subjective theoretical framework derived from expert opinion [43]. Alternatively, summaries with specific weightings can be used, such as the General index [44] and the geoindex mean of multiple relative abundance indices (often used to quantify biodiversity [45]). A fourth approach is to obtain composite indices using Principal Component Analysis (PCA). This follows a more data-driven approach to combine multiple indices into a single real-valued score. These composite indices are commonly used for estimating general indicators of ecological systems [46], such as ecological integrity [47] and multispecies biodiversity indicators [48], rather than abundance. These methods do not respect any of the criteria C1 to C4, and C6 [43].

Geostatistical methods have been developed for modelling multiple indices of animal abundance for a single species. However, these methods were found to either use one index as a predictor for another index [49], thus violating C1 and C2, or to use multivariate kriging for all indices [50], violating C1 and C6.

There are several examples of geostatistical approaches which jointly model indices for multiple species in the field of ecological community modelling [42, 51, 52]. However, the structure of these models does not enable predictions to be made for the abundance of a single species measured by multiple indices, as is required for C4. They were also found to require all indices to be measured at a common set of locations, hence violating C2. While integrated species distribution models (ISDMs) offer a means to model multiple indices, they have been developed to combine multiple presence-only and presence-absence data sources [42, 53], rather than abundance indices. These models also provide no way to explore the relative contributions of each data source. Consequently, they do not meet C4 and C5.

### 2.2 Model formulation and inference

Let *R*(*x*) denote a spatially continuous stochastic process, representing *rattiness*, our proxy for rat abundance. The data consist of a set of outcomes *Y*_*i*_ = (*Y*_*i,j*_ : *j* = 1, …, *J*), for *i* = 1, …, *N*, collected at a discrete set of locations *X* = {*x*_*i*_ : *i* = 1, …, *N*}. The outcome variables *Y*_*j*_ : *j* = 1, …, *J* are a set of indices that provide information about *R*(*x*).

Let “[*·*]” be a shorthand notation for “the probability distribution of *·*.” Define *Y* = (*Y*_1_, …, *Y*_*N*_) and *R* = (*R*(*x*_1_), …, *R*(*x*_*N*_)). We assume that the *Y*_*i,j*_ : *j* = 1, …, *J* are conditionally independent given *R*(*x*_*i*_), as formally expressed by

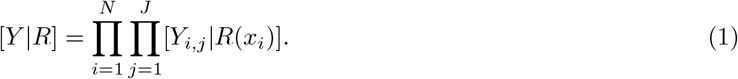

Let *g*_*j*_(*·*) and *η*_*j*_(*x*_*i*_) denote the link function and linear predictor for the outcome variables *Y*_*i,j*_ : *i* = 1, …, *N*. Hence,

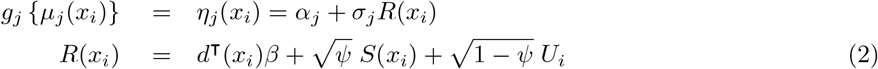

where: *d*(*x*_*i*_) is a vector of explanatory variables with associated regression coefficients *β*; *U*_*i*_ is a set of independently and identically distributed zero-mean Gaussian variables with unit variance; *S*(*x*_*i*_) is a stationary and isotropic spatial Gaussian process; *σ*_*j*_ > 0 : *j* = 1, …, *J* are scale parameters that account for the different scales of variation of the linear predictors of each outcome *Y*_*i,j*_; *ψ ∈* (0, 1) regulates the relative contributions of spatially structured variation, *S*(*x*_*i*_), and unstructured random variation, *U*_*i*_, to *R*(*x*_*i*_).

For the analysis of Section 3, we specify an exponential spatial correlation function

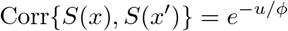

where *u* = ||*x* –*x′*|| is the Euclidean distance between *x* and *x′*, and *ϕ* regulates how fast the spatial correlation decays to zero with increasing distance *u*.

To fit the model in (2), we use the Monte Carlo maximum likelihood (MCML) method [54] and proceed as follows. Let *θ* = (*α*_1_, …, *α*_*J*_, *σ*_1_, …, *σ*_*J*_) and *ω* = (*β, ϕ, ψ*) be the vector of unknown parameters associated with [*R*] and [*Y* |*R*]. The likelihood function is then given by

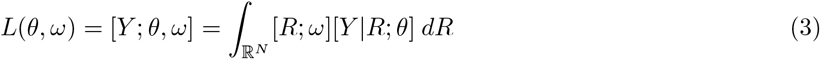

Since the integral in (3) cannot be solved analytically, we approximate it using Monte Carlo methods. Specifically, let *θ*_0_ and *ω*_0_ be our initial best guesses for *θ* and *ω*, respectively. Since [*R*; *ω*][*Y* |*R*; *θ*] *∝* [*R*|*Y*; *ω*] we re-write the integral in (3) using an importance sampling distribution [*R*; *ω*_0_][*Y* |*R*; *θ*_0_] to give

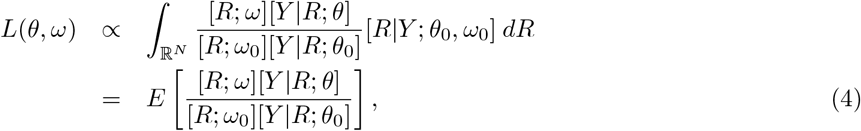

where the expectation is taken with respect to the distribution of [*R*|*Y*; *ω*_0_].

Based on (4), we then approximate (3) with

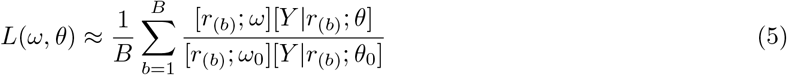

where *r*_(*b*)_ is the *b*-th sample from [*R*|*Y*; *ω*_0_,*.θ*_0_]. To obtain the maximum likelihood estimate for *θ* and *ω*, we maximize (5) using numerical optimization. To simulate from [*R*| *Y*; *θ*_0_, *ω*_0_], we use the Laplace sampling algorithm described in detail in [54] and [55].

To improve the approximation of the likelihood function, we also update our guesses *ω*_0_ and *θ*_0_ by plugging in the maximum likelihood estimate and re-iterate the maximization of (5) until convergence.

### 2.3 Exploratory analysis

In this section we outline several key steps in an exploratory analysis of the data to guide the model-building process.

The exploratory analysis serves three purposes: i) to explore the relationship between the latent *rattiness* process, *R*(*x*), and the covariates *d*(*x*); ii) to test for the presence of residual spatial variation in *R*(*x*) unexplained by the covariates *d*(*x*); and iii) to assess if the data support the assumed stochastic dependence structure as represented by the causal arrows of Figure 1.

**Figure 1:**
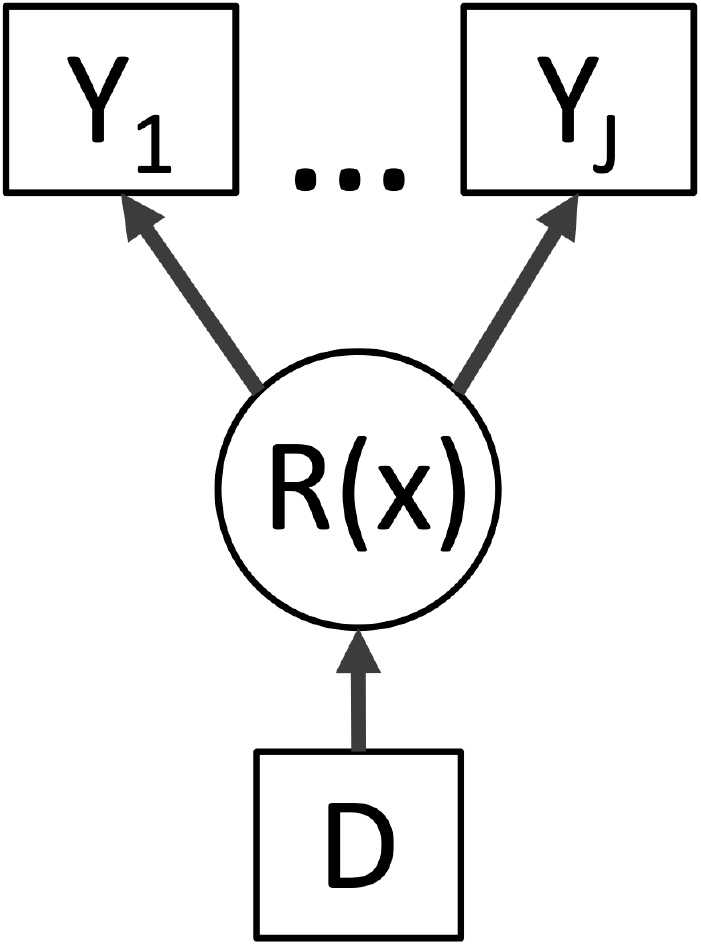
Directed acyclic graph (DAG) of the geostatistical model of Section 2.2. R(x) is the value of a spatially continuous stochastic process at location x. The outcome variables Y_j_ : j = 1, …, J are a set of indices that provide information about R(x). The term D represents a set of explanatory variables which contribute to the spatial variation in R(x). Square objects correspond to observable variables, and circles to latent random variables.

To pursue i), we first analyse the data using a simplified version of the model in (2) that does not assume spatial correlation and does not make use of any of the available covariates by setting *ψ* = 0 and *β* = 0. Rattiness is consequently modelled purely as unstructured random variation, hence *R*(*x*_*i*_) = *U*_*i*_. Note that the likelihood associated with *θ* = (*α*_1_, …, *α*_*J*_, *σ*_1_, …, *σ*_*J*_) and *ω* = (*ϕ*), is now given by the product of *N* one-dimensional integrals,

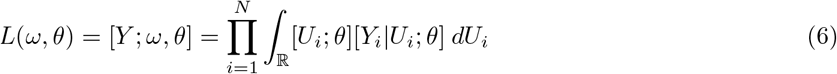

To maximize the likelihood, each of the factors in the above product can then be approximated using numerical quadrature; we use a quasi Monte Carlo method whereby the integrals in (2) are drawn deterministically based on the Halton sequence of support points [56].

After fitting the model in (6), we then estimate *U*_*i*_ using its predictive expectation,

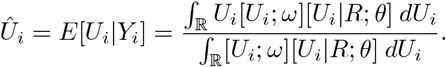

To compute the *Û*_*i*_ we plug in the maximum likelihood estimates for *θ* and *ω*. To explore the functional form of the relationship between *R*(*x*_*i*_) and the explanatory variables *d*(*x*_*i*_), we plot the *Û*_*i*_ against the values of *d*(*x*_*i*_). Based on the empirical relationship observed in the scatter-plots, we then introduce *d*(*x*_*i*_) into the linear predictor for *R*(*x*_*i*_), leading to a first extension of the simplified model to

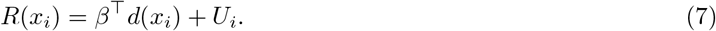

We can now use the model in (7) to pursue the second objective, i.e. testing for residual spatial correlation. We then re-fit the likelihood (6), without setting *β* to zero, and re-compute *Û*_*i*_, which now represent our provisional estimate for the residual variation in *R*(*x*_*i*_) that is unexplained by *d*(*x*_*i*_). To check if the *Û*_*i*_ show evidence of spatial correlation, we then randomly permute locations *x*_*i*_ in the data while holding the *U*_*i*_ fixed and repeat this 10,000 times. For each of the permuted data-sets, we compute the empirical variogram based on the *Û*_*i*_ and use the resulting 10,000 variograms to compute 95% confidence intervals under the assumption that the *Û*_*i*_ are not spatially correlated. If the variogram computed from the original *Û*_*i*_ falls fully within the 95% band, we conclude that the *Û*_*i*_ do not show evidence of residual spatial correlation. If the variogram partly falls outside the 95% band, we conclude that there is evidence of spatial correlation and fit the geostatistical model defined in Section 2.2.

Finally, for the third objective, we test the null hypotheses *H*_0_ : *σ*_*j*_ = 0, for *j* = 1, …, *J* using likelihood ratio tests. Note that, in this case, the conditions that give an asymptotic distribution of the likelihood ratio test based on a *χ*^2^ distribution are not met because the value 0 is on the boundary of the parametric space of *σ*_*j*_; following [57], we correct the nominal resulting p-values by multiplying them by 1*/*2.

### 2.4 Spatial prediction and assessment of the contribution of each index of abundance

Our predictive target is 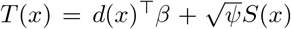 at a set of prediction locations, 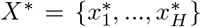. To predict 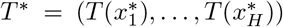, we sample from its predictive distribution [*T* ^∗^|*Y*] as follows. We first simulate from [*R*|*Y* ; *θ, ω*] using the same sampling algorithm as for maximizing the likelihood in Section 2.2, with the parameters *θ* and *ω* fixed at their maximum likelihood estimates. After obtaining samples *r*_(*b*)_, *b* = 1, …, *B*, we then simulate from [*T* ^∗^|*r*_(*b*)_], which follow a multivariate Gaussian distribution with mean and covariance matrix easily obtained from their joint Gaussian distribution [*R, T*^∗^]. From this, we obtain 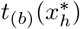, for *h* = 1, …, *H* and *b* = 1, …, *B*, which are now samples drawn from [*T* ^∗^|*Y*]. These can be used to compute any desired summary of the predictive distribution [*T* ^∗^*|Y*], such that the expectation

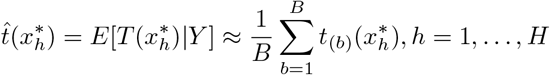

and the standard deviation

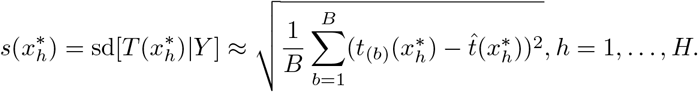

To assess the contribution of each index to the prediction target *T*^∗^, we then compare 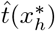 and 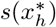 from the geostatistical model fitted using all indices with those obtained from *J* models, each of which excludes a single index.

Let 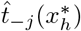 and 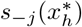 denote the predictive mean and standard deviation obtained from excluding data for the *j*-th index; to summarize the discrepancy between this and the full model, we average the squared differences across all locations *X*\*, i.e.

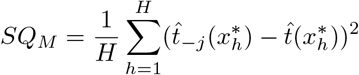

and

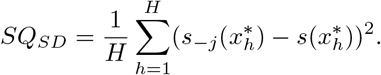

## 3 Case study: Norway rats in Pau da Lima community

### 3.1 Study site and data collection

Pau da Lima (13°32’53.47” S; 38°43’51.10” W) is a low-income urban community with a high annual leptospiral infection rate of 35.4 (95% CI, 30.7–40.6) infection events per 1,000 annual follow-up events in the period 2003 to 2007 [32, 58]. It is located on the periphery of the city of Salvador in Northeast Brazil and has been a focus for leptospirosis research for over fifteen years [30, 32, 41]. The study site at Pau da Lima is characterised by three valleys (see Figure 2) with large elevation gradients, high population density, poverty, low levels of education and poor provision of sanitation and refuse collection services.

**Figure 2:**
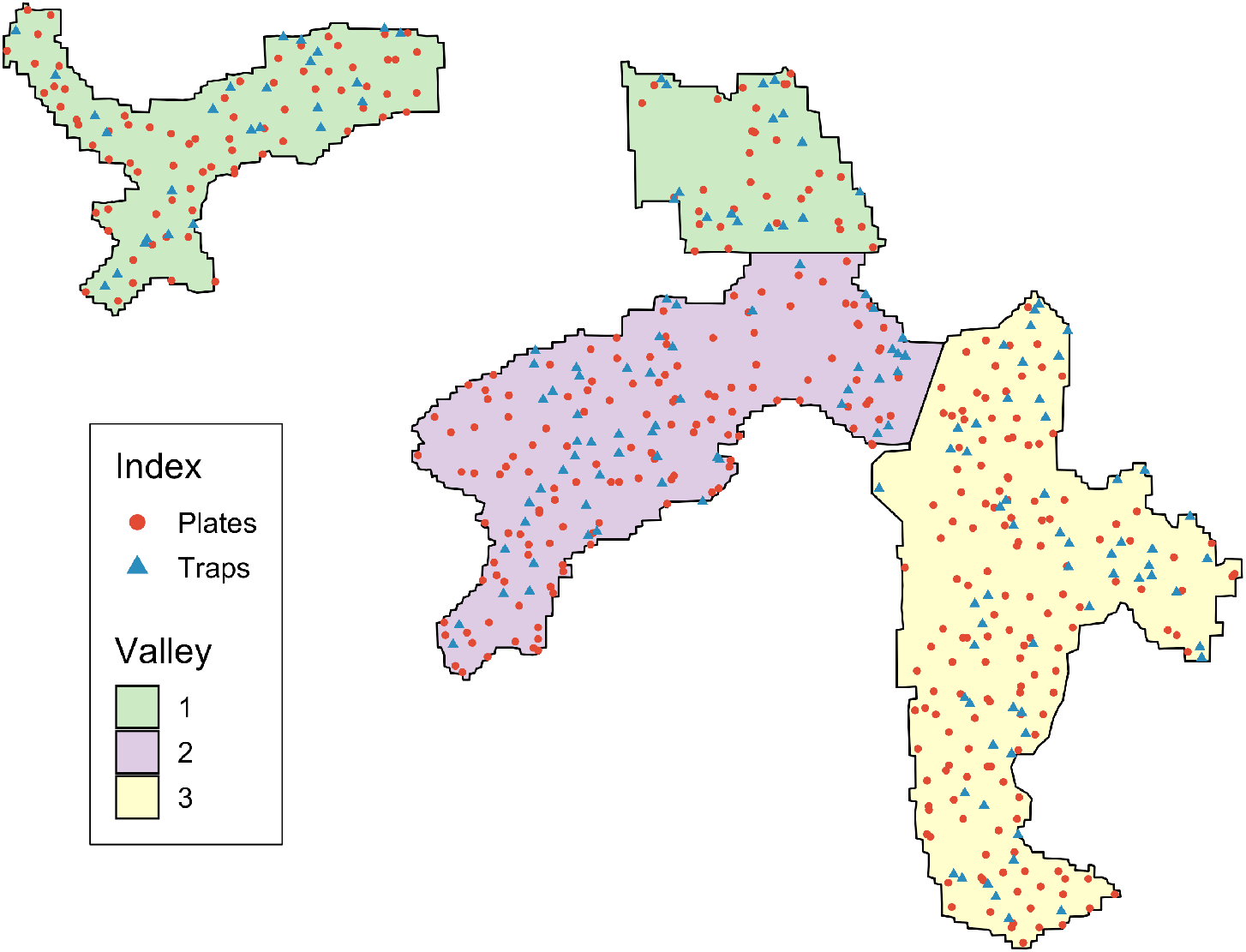
Map of the three valleys within the study site, Pau da Lima, with sampled locations for track plates and traps shown. Surveys for signs of rat infestation were conducted at all locations.

To describe the spatial variation in rat abundance within Pau da Lima, a cross-sectional study was conducted from October to December 2014. Rat trapping was carried out at 159 locations across the study area with two traps deployed for 4 consecutive 24-hour trapping periods at each point (see Panti-May *et al*. [41]). After each 24-hour period, trapping success and closure of the trap without a rat caught inside were recorded. Track plates were used for two consecutive 24-hour periods at 415 locations [59] using the standardised protocol for placement and survey developed and validated previously [6] with five plates placed at each location in the shape of a ‘five’ on a die with 1 metre spacing between each plate. After each 24-hour period, plates were repainted and lost plates were recorded and replaced. A map of the study area and sampling locations for rat trapping and track plates is shown in Figure 2. A survey for signs of rat infestation (presence of trails, faecal droppings and active burrows), adapted from the Centers for Disease Control and Prevention [60] and validated in the study area [34], was also conducted at all locations at which traps or track plates were deployed. In our analysis, we consider the following environmental variables: elevation, distance to public refuse dumps and the proportion of land cover classified as vegetation within a 30 metre radius. Land cover data was created by classification of Digital Globe’s WorldView-2 satellite imagery (8 bands with resolution 0.5m by 0.5m resolution taken on February 17, 2013) [61] using a maximum likelihood supervised algorithm. This was validated with ground truthing data collected from 20 randomly selected sites of size 5m by 5m.

### 3.2 Applying the model to Pau da Lima data

In this analysis, we shall also refer to *R*(*x*) as the *rattiness* process. To make inference on *R*(*x*), we shall use data collected on three indices: rat signs (*j* = 1), live traps (*j* = 2) and track plates (*j* = 3).

Due to theft, violence and heavy rainfall, 993 out of 4150 (23.9%) plates were lost, with all five plates lost at 126 out of 830 (15.2%) track plate days. For the traps, 85 out of 1272 (6.7%) trapping-days were lost, of which 458 (38.6%) were found closed and empty after a 24-hour period. This is a common issue and is due to traps malfunctioning or being tampered with by animals or people. Of the remaining track plate-days, 263 (37.4%) had at least one plate with rat markings. Similarly, 200 (34.8%) of surveys found at least one sign of rat infestation. Of the trapping-days which were not lost, the trapping success rate was low, with only 112 (9.4%) trapping-days found to have caught a rat.

Let the variable *Y*_*i*,1_ be a binary indicator taking value 1, if at least one sign of rat infestation was found at location *x*_*i*_ and 0 otherwise. We model the probability of finding a sign of rat infestation, *µ*_1_(*x*_*i*_), using a logit-linear regression log{*µ*_1_(*x*_*i*_)*/*(1 − *µ*_1_(*x*_*i*_))} = *α*_1_ + *σ*_1_*R*(*x*_*i*_)

The variable *Y*_*i*,2_, conditionally on *R*(*x*_*i*_), is a Binomial variable representing the number of traps, out of *n*_*i*,1_, in which rats were captured. We assume that the times of rat captures from a trap follow a time-varying inhomogeneous Poisson process with intensity *t*_*i*_*µ*_2_(*x*_*i*_), where *t*_*i*_ is the time (in days) for which a trap is operative and log{*µ*_2_(*x*_*i*_)} = *α*_2_ + *σ*_2_*R*(*x*_*i*_). It follows that the probability of capturing a rat is

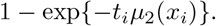

If a trap is found closed without a rat, we assume that the trap was disturbed and set *t* = 0.5. In all other cases, we set *t* = 1.

Finally, *Y*_*i*,3_, is the number of track-plates, out of *n*_*i*,3_, that show presence of rats. We model this as a Binomial variable with *n*_*i*,3_ trials and probability *µ*_3_(*x*_*i*_) where log{*µ*_3_(*x*_*i*_)*/*(1 − *µ*_3_(*x*_*i*_)(*x*_*i*_)} = *α*_3_ + *σ*_3_*R*(*x*_*i*_).

## 4 Results

### 4.1 Exploratory analysis

Following the steps outlined in Section 2.3, we first fit the simplified non-spatial model (*R*(*x*_*i*_) = *U*_*i*_) to explore the relationship between *rattiness* and each of the environmental variables considered. The results are reported in the scatter-plots of Figure 3. In Figure 3a, we observe a negative linear relationship between elevation and *rattiness*. In Figure 3b, we notice that, on average, higher values of *rattiness* are observed for distances from dumps less than 90 meters. Finally, Figure 3c, shows that the mean proportion of land cover classified as vegetation (within a 30 meters radius of a given sampling point), is approximately linearly and positively associated with *rattiness*.

**Figure 3:**
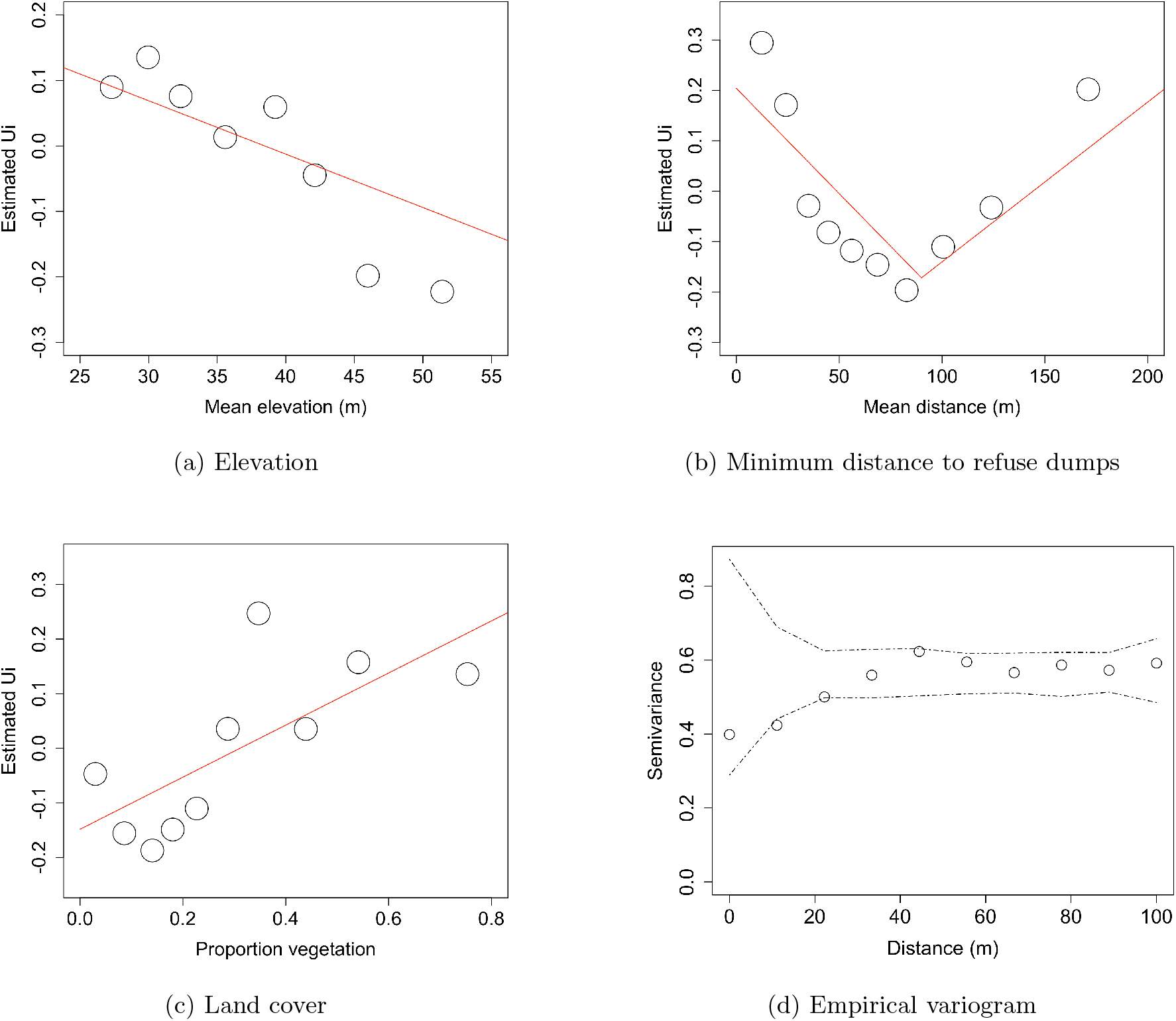
Panels (a) to (c) are scatter-plots of the unstructured random variation, Û_i_, against the covariates considered in the analysis. Û_i_ are estimated using a non-spatial model which excludes all covariates. The relationship between each covariate and Û_i_ is shown with a red line and was estimated using univariable linear regression (with a linear piecewise spline knot included at 90 metres for the distance to refuse covariate). Panel (d) is a variogram computed from Û_i_ using a non-spatial model that includes all of the covariates; the dashed lines correspond to 95% confidence intervals under the assumption of spatial independence.

Based on the results of this exploratory analysis, we then extend the non-spatial model to include the covariate effects on *rattiness*. Hence, our model for *R*(*x*_*i*_) becomes

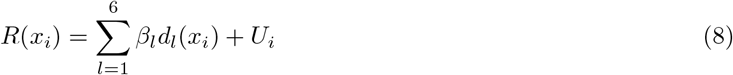

where each of the terms *β*_*l*_ corresponds to covariate effects as defined in Table 1, *d*_*l*_ are the explanatory variables and *U*_*i*_ are the unstructured random effects.

**Table 1:** Regression coefficients of the environmental covariates used to model *rattiness*.

| Term | Variable |
| --- | --- |
| $\beta_1$ | Elevation (m) |
| $\beta_2$ | Refuse distance (0-90m) |
| $\beta_3$ | Refuse distance (>90m) |
| $\beta_4$ | Proportion of land cover vegetation (%) |
| $\beta_5$ | Valley 2 (relative to Valley 1) |
| $\beta_6$ | Valley 3 (relative to Valley 1) |

We then carry out the likelihood ratio tests for testing the three hypothesis *H*_0_ : *σ*_*j*_ = 0 for *j* = 1, 2, 3. All three yield p-values less than 0.0001, supporting the use of a joint model for all three indices.

Figure 3d shows the variogram for the *Û*_*i*_ obtained from the spatially uncorrelated model (8). Most of the points of the empirical variogram lie inside the 95% tolerance band but the variogram point at around 10 meters is a highly unlikely value under the assumption of spatial independence. Since the variogram diagnostic does not provide an unequivocal answer to the question of whether a spatially correlated term is needed, we fit a geostatistical model to assess this.

### 4.2 Geostatistical model

The parameter estimates based on the Monte Carlo maximum likelihood method are reported in Table 2 under the ‘full model’ column. The estimate for the scale of spatial correlation, *ϕ*, of about 13 meters indicates that the data exhibit spatial correlation after controlling for the explanatory variables. The estimate for *ψ* of about 0.9 implies a greater contribution to *rattiness* from the Gaussian process, *S*(*x*_*i*_), than from the unstructured random variation, *U*_*i*_, suggesting that most of the unexplained variation in *rattiness* is spatially structured. All the point estimates of the regression coefficients *β*_*l*_ are consistent with the scatter-plots of Figure 3. Valleys 2 and 3 both had lower mean levels of *rattiness* relative to Valley 1, controlling for all other covariates. While the association with valley was significant at *p <* 0.05, elevation, distance to refuse dumps or land cover covariates were not significant.

**Table 2:** Parameter estimates for the full model and the three two-indices models where *α*_1_, *α*_2_ and *α*_3_ (and *σ*_1_, *σ*_2_ and *σ*_3_) denote the coefficients for Signs, Traps and Plates, respectively

| Parameter | Estimate (95%CI) |  |  |  |
| --- | --- | --- | --- | --- |
|  | Full model | Signs & Traps | Traps & Plates | Signs & Plates |
| $\alpha_1$ | -0.642 (-0.891, -0.425) | -0.508 (-0.997, -0.115) | - | -0.630 (-0.853, -0.419) |
| $\alpha_2$ | -2.684 (-3.078, -2.361) | -2.607 (-3.021, -2.099) | -2.456 (-2.849, -2.127) | - |
| $\alpha_3$ | -2.503 (-2.925, -2.144) | - | -2.567 (-3.066, -2.119) | -2.672 (-3.090, -2.305) |
| $\sigma_1$ | 0.747 (0.455, 1.045) | 1.040 (0.615, 1.356) | - | 0.641 (0.372, 0.906) |
| $\sigma_2$ | 0.920 (0.613, 1.183) | 1.068 (0.546, 1.182) | 0.863 (0.436, 1.133) | - |
| $\sigma_3$ | 1.896 (1.557, 2.179) | - | 1.795 (1.348, 2.174) | 1.877 (1.542, 2.163) |
| $\phi$ | 13.432 (6.833, 21.172) | 46.413 (7.692, 162.455) | 11.306 (4.978, 17.325) | 12.270 (6.920, 16.099) |
| $\psi$ | 0.878 (0.529, 1.000) | 0.492 (0.161, 0.878) | - | - |
| $\beta_1$ | -0.131 (-0.333, 0.058) | -0.337 (-0.795, -0.050) | -0.103 (-0.396, 0.158) | -0.164 (-0.370, 0.024) |
| $\beta_2$ | -0.234 (-0.582, 0.101) | -0.173 (-0.819, 0.407) | -0.299 (-0.795, 0.175) | -0.055 (-0.401, 0.295) |
| $\beta_3$ | -0.074 (-0.399, 0.260) | 0.016 (-0.343, 0.354) | -0.129 (-0.493, 0.077) | -0.011 (-0.125, 0.248) |
| $\beta_4$ | 0.114 (-0.075, 0.310) | 0.158 (-0.170, 0.527) | 0.086 (-0.194, 0.376) | 0.115 (-0.070, 0.308) |
| $\beta_5$ | -0.229 (-0.358, -0.108) | 0.116 (-0.170, 0.354) | -0.269 (-0.485, -0.085) | -0.361 (-0.503, -0.241) |
| $\beta_6$ | -0.159 (-0.289, -0.034) | 0.195 (-0.064, 0.459) | -0.220 (-0.401, -0.040) | -0.182 (-0.307, -0.059) |

### 4.3 Spatial prediction

Our predictive target for *rattiness* is

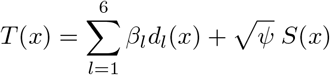

for prediction locations *x* forming a 5 m by 5 m regular grid covering the whole of the study area.

Maps for the mean and standard deviation of the predictive distribution of *T* (*x*) are shown in Figure 4. These show a highly heterogeneous spatial pattern with localized hotspots in Valley 1 (Figure 4a) and in the low elevation central regions of the two other valleys. Areas with low values of predicted *rattiness* in Valleys 2 and 3 are characterized by a high proportion of soil land cover and higher elevations. The Gaussian process, *S*(*x*), contributed more than the covariates to the prediction of the *rattiness* surface shown in Figure 4b. This is evidenced by the clear geographical overlap of hotspots in both *S*(*x*) and mean predicted *rattiness*. As expected, in areas with fewer or no observations, standard errors are larger than elsewhere.

**Figure 4:**
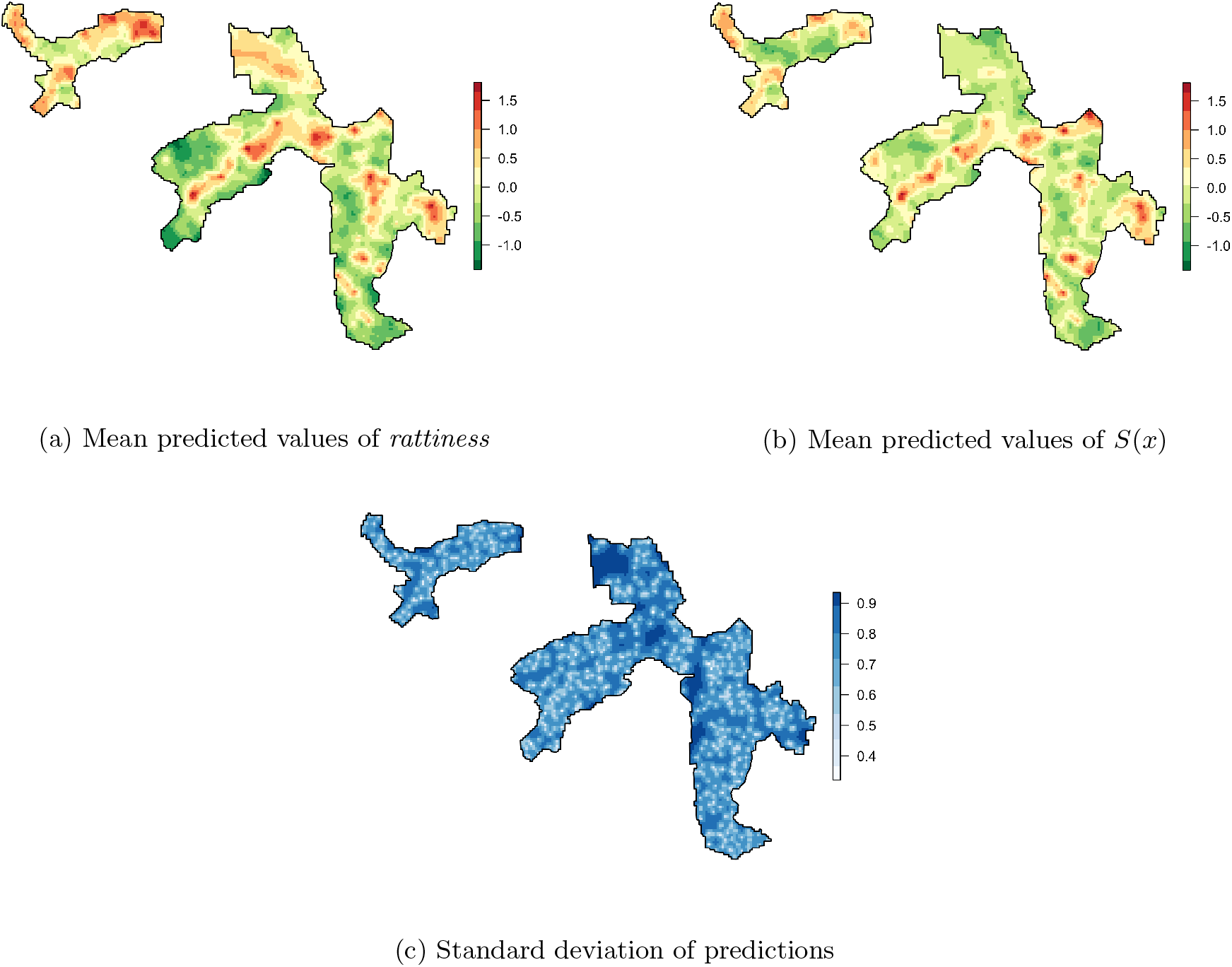
Rattiness model predictions (positive rattiness values indicate areas of high relative abundance)

### 4.4 Relative contributions of indices

To assess the impact of each index on parameter estimation, we fitted three models, each discarding one of the three indices; the parameter estimates are reported in Table 2, and are similar to those estimated for the full model. Across all four models, the signs of covariate estimates were consistent except for the valley indicator variables, which vary both in sign and magnitude. The estimates for the scale of the spatial correlation, *ϕ*, from the “Traps & Plates” and “Signs & Plates” models are close to that of the full model, but the “Signs & Traps” model had a substantially larger estimate of about 46 metres with a much wider confidence interval. The increased uncertainty in the estimation of the spatial correlation after excluding plates suggests that this index may be one of the main factors driving our predictions for *rattiness*. Furthermore, when the plates are included in the model, the spatial variation entirely dominates the *rattiness* process with estimates for *ψ* very close to 1. We therefore fixed *ψ* = 1 for the “Track & Plates” and “Sign & Plates” models.

To visualize the differences in the spatial predictions for *rattiness* and *S*(*x*), we compute the relative difference between the predictions obtained from each of the models excluding one of the three indices and the full model. Figure 5 reports the maps of the relative differences for *rattiness*, and Figure 6 for *S*(*x*). The spatial predictions from the “Traps & Plates” and “Signs & Plates” models were more similar to those obtained from the full model as indicated by relative differences close to zero throughout the study area in both figures. In contrast, the predictions for the “Signs & Traps” model were different to those made by the three other models in most parts of the study area, with relative differences ranging from about −2 to +1.

**Figure 5:**
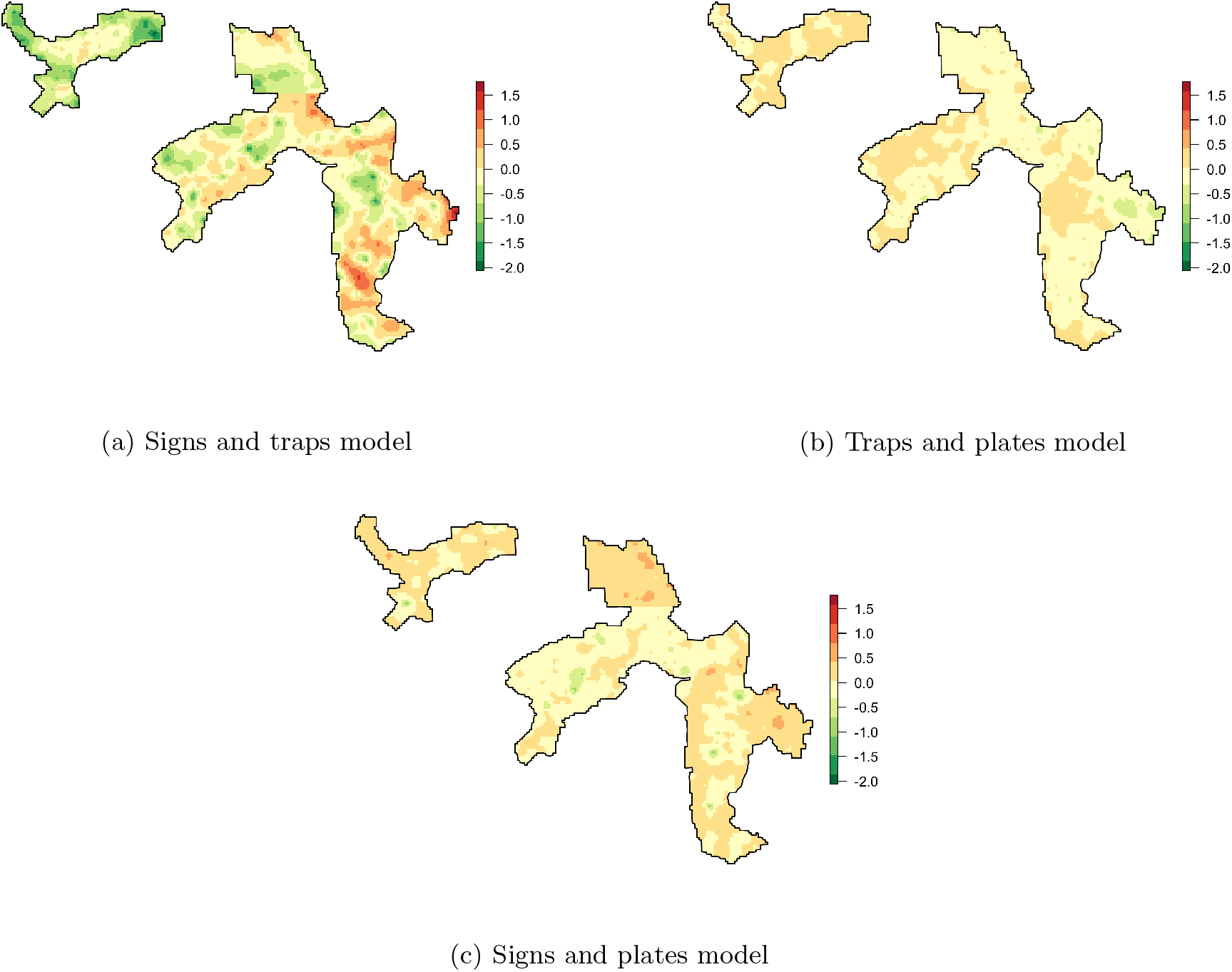
Model predictions for rattiness for each two-indices model relative to the full model (a value of zero indicates that there is no difference in the models’ predicted values)

**Figure 6:**
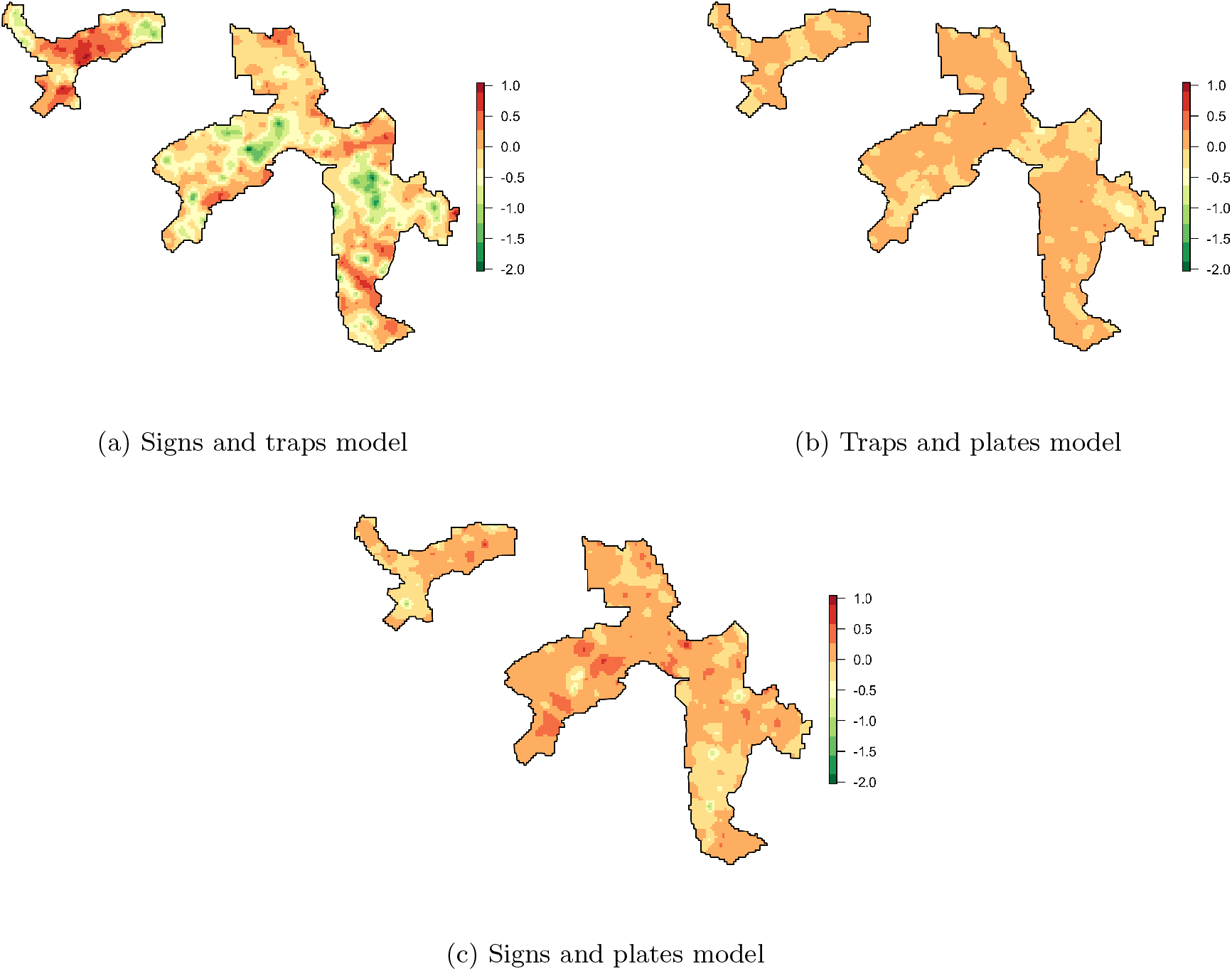
Model predictions for the spatial Gaussian process, S(x), for each two-indices model relative to the full model (a value of zero indicates that there is no difference in the models’ predicted S(x) values)

Table 3 reports the *SQ*_*M*_ and *SQ*_*SD*_ summaries used to quantify the changes in point predictions and standard deviations for *rattiness*; see Section 2.4 for a formal definition of these two summaries. The results clearly highlight the model excluding plates as yielding substantially different predictions for *rattiness*, as well as larger standard errors. These results are consistent with our findings from Table 2, providing further evidence on the importance of plates to our predictive inferences on *rattiness*.

**Table 3:** The squared differences of the point predictions (*SQ*_*M*_) and standard deviations (*SQ*_*SD*_) between the full model and each of the three two-indices models, averaged over all the prediction locations. For a formal definition of *SQ*_*M*_ and *SQ*_*SD*_, see Section 2.4.

| Model | $SQ_M$ | $SQ_{SD}$ |
| --- | --- | --- |
| Traps & Plates | 0.018 | 0.007 |
| Signs & Plates | 0.032 | 0.003 |
| Signs & Traps | 0.249 | 0.096 |

## 5 Discussion

In this paper, we have developed a flexible geostatistical framework that borrows information across multiple indices of vector and reservoir host abundance to carry out spatial prediction for a shared latent variable that acts as a proxy for animal abundance. To our knowledge this is the first study that proposes a multivariate geostatistical framework to jointly model multiple indices of abundance for a single species using a statistically principled likelihood-based approach.

We have applied the method to mapping *rattiness*, a proxy for Norway rat abundance, in a low-income urban community in Salvador, Brazil. We found that *rattiness* is lower at higher elevations and longer distances from large refuse piles, and higher in more densely vegetated areas. In our study site, elevation was used as a proxy for socioeconomic status within the community, with improved housing quality, sanitary conditions and road surfacing found at higher elevations. The point estimates for the regression coefficients associated with each of these variables are consistent with previous studies [37, 62], and can be explained by Norway rats’ preference for habitats with greater access to food and harbourage. However, the inherently high sampling variation in the data recorded by each of the three indices results in wide confidence intervals for these estimates. A separate analysis that included elevation as the only covariate for *rattiness* (Supplementary Material 1) supports the conclusion that this was a key source of uncertainty; the effect of elevation was statistically significant at the conventional 5% level. Measurement error in covariates is also a likely contributor to these wide confidence intervals. This is a common issue when prediction is the priority, as was the case in our application, and covariates are constrained to those for which values are available at all prediction locations.

A strength of our model is its ability to borrow information across space without requiring data from multiple indices to be co-located. This is especially useful when combining data for multiple indices from separate studies, or when there is a non-negligible loss or malfunctioning of measurement tools [63]. This arose in our analysis of the Norway rat due to track-plate loss and empty closing of traps, and is commonly encountered when measuring rodent abundance [2]. The framework also provides flexibility for modelling indices of abundance that are measured on a wide range of different scales with different sampling distributions. This was useful in our application as we were able to use a non-standard approach to more accurately model the trapping process and account for trap failure.

The estimate for the spatial correlation parameter, 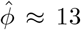 metres corresponds to a spatial correlation range (the distance at which the correlation reduces to 0.05) of approximately 40 metres. This is consistent with studies investigating the size of the main activity area of Norway rats, which has been estimated to have a radius of 25–150m in urban areas [64, 65]. In environmentally heterogeneous and resource-rich areas, such as Pau da Lima, the size of a rat’s activity space has been found to be smaller, as shown by estimates of population density varying significantly within a city block [66] or along the length of an alley [67]. This is due to strong spatial heterogeneity in the presence of food and harbourage, availability of access routes and the presence of barriers to movement. All of these can result in high site fidelity (a measure of how concentrated an animal’s movements are around a specific site) [66] and significant variations in the abundance and activity of rats over small distances. The estimated value of *ψ* indicates that the spatially structured random effects are more important than the non-structured random effects in predicting *rattiness*. This follows from the fact that they account for unmeasured variables of habitat suitability, which can be expected to be spatially structured.

The finding that track plates were an important contributor for *rattiness* estimation indicates the greater information content provided by this tool relative to traps and signs. Nevertheless, the other two indices also contributed to *rattiness*, enabling more precise predictive inferences to be made than could be obtained using only the track-plates data. Due to the scarcity of resources available for monitoring programmes of vector and animal reservoir populations, efficient data collection is critical, making the choice of which indices, or how many should be used, an important consideration. A key strength of our methodology is that it provides the user with the tools to explore the contributions of each index to *rattiness* (or the spatial latent process for any other application). The likelihood-ratio tests described in Section 2.3 can be used to check which indices contribute to the model for prediction of rattiness. Any two such indices must both be associated with the latent *rattiness* variable, R(x), in Figure /1, and therefore necessarily with each other. However, if two indices are near-collinear the likelihood-ratio test will indicate that one of the two is redundant. The *SQ*_*M*_ and *SQ*_*SD*_ summaries then enable the user to evaluate the extent to which each included index contributes to the predictions.

The interpretation of the *rattiness* process is an important, context-dependent issue. For example, if all the indices used in the analysis are reliable indicators of abundance, then *rattiness* has a clear interpretation as an overall measure of abundance. However, tools used to estimate abundance often provide measures that are a mixture of both animal abundance and behaviour [3]. In our application, the rat signs are an index of rat presence, while track plates and traps both measure abundance and activity, with track plates more strongly representative of activity. The resulting measure of *rattiness* obtained by combining the three indices therefore represents a data-determined synthesis of these three processes. In our motivating example concerning the role of rats in determining risk of human leptospirosis, the mechanism though which the vector confers disease risk is driven by both the size of the rat population and its behaviour, of which activity is one aspect [68, 69]. For this reason, we argue that joint modelling of multiple indices can be especially relevant to understanding geographical variation in disease risk.

The proposed modelling framework can also be applied to problems of environmental management and conservation, where indices of abundance are widely used to monitor wild animal populations and their impact on biodiversity, agriculture or another species’ ability to survive, and to guide management decisions [3, 63, 70]. Examples of recent studies that have used multiple indices of abundance include: invasive roof rat and deer mouse populations in orchards [71]; the threatened survival of native species due to invasive small mammals on islands [72]; the impact of rats and possums in New Zealand [73]; and, the effect of elephants on woody vegetation in sub-Saharan Africa [74].

One limitation of our approach is that it does not account for detection bias. Methods that account for this bias require absolute abundance data, which is difficult to collect, or data collected using double-sampling techniques that require an often unattainable trapping success rate - for rodents, of at least 20% [2, 75]. For this reason, index data is still widely collected for monitoring purposes without the requirement for these additional data sources.

## Data Availability

R scripts for the rattiness model and all data needed for our analysis are available at https://github.com/maxeyre/Rattiness-1.

## Notes

### Competing Interest Statement

The authors have declared no competing interest.

### Funding Statement

Funding for this project was received from: the Minist é
rio da Ciência, Tecnologia e Inovaçâo, Brazil through the Conselho Nacional de Desenvolvimento Científico e Tecnológico and
Fundaçâo de Amparo à Pesquisa do Estado da Bahia, Secretaria Municipal de Saúde de Salvador-BA, Fundaçâo Oswaldo Cruz (grant 10206/20155), Medical Research Council, UK (grants 1964635, MR/P024084/1, MR/T029781/1), the Ministério da Saúde, Brazil, the National Institute of Allergy and Infectious Diseases, USA (grants F31 AI114245, R01 AI052473, R01 TW009504, R25 TW009338, U01 AI088752) and the Wellcome Trust (grants 102330/Z/13/Z, 218987/Z/19/Z).

### Author Declarations

The ethics committee for the use of animals from the Oswaldo Cruz Foundation, Salvador, Brazil approved the protocols used in this study (protocol number 003/2012), which adhered to the guidelines of the American Society of Mammalogists for the use of wild mammals in research (Sikes and Gannon, 2011) and the guidelines of the American Veterinary Medical Association for the euthanasia of animals (Leary et al., 2013). These protocols were also approved by the Yale University Institutional Animal Care and Use Committee (IACUC), New Haven, Connecticut (protocol number 2012-11498). Sikes RS, Gannon WL, Animal Care and Use Committee of the American Society of Mammalogists. Guidelines of the American Society of Mammalogists for the use of wild mammals in research. J Mammal. 2011; 92:235-253. Leary S, Underwood W, Lilly E, Anthony R, Cartner S, Corey D, et al. AVMA guidelines for the euthanasia of animals: 2013 Edition. Schaumburg, Illinois: American Veterinary Medical Association; 2013.

